# Mortality in Rural and Urban Emergency and Trauma Care in the United States: A Systematic Review and Meta-Analysis

**DOI:** 10.64898/2026.10.12.26365329

**Authors:** Ahmed Elbasty, Ranjeet Sunkara, Madhav Arora, Tehreem-Un-Nisa, Vivin Mohankumar

**Author notes:** Corresponding author: Ahmed Elbasty.

## Abstract

**Objective:** Rural hospitals in the United States have fewer specialists, fewer critical care resources, and longer transport distances than urban hospitals, yet national studies disagree on whether rural patients are more likely to die after emergency care. The goal of this study was to determine whether short-term mortality differs between rural and urban patients receiving emergency or trauma care and to estimate the size of the difference.

**Methods:** We conducted a systematic review and meta-analysis reported in line with PRISMA 2020. PubMed was searched on October 11, 2026, for studies published from 2010 onward, supplemented by web searching. Eligible studies were US observational studies that compared rural with urban patients or hospitals and reported mortality after emergency or trauma care. Odds ratios (ORs) were pooled with a DerSimonian-Laird random-effects model, with Hartung-Knapp confidence intervals (CIs) as a sensitivity analysis. Injury and trauma studies formed the primary analysis. Certainty of evidence was rated with GRADE.

**Results:** Of 161 records identified, 14 studies were included and 12 were pooled. Across seven injury and trauma studies, rural setting was associated with higher odds of death (OR, 1.27; 95% CI, 1.07-1.52), with substantial heterogeneity (I^2^ = 84%; 95% prediction interval, 0.74-2.19). Across all 12 studies, the pooled OR was 1.23 (95% CI, 1.09-1.40; I^2^ = 96%). With Hartung-Knapp intervals, the overall estimate remained significant (95% CI, 1.05-1.45) and the trauma estimate was borderline (95% CI, 0.99-1.64). Study estimates ranged from 0.65 to 1.97. Three pediatric trauma studies gave a consistent estimate (OR, 1.32; 95% CI, 1.18-1.47; I^2^ = 0%). Subgroup analyses by rurality definition, age group, and geographic scope did not explain the heterogeneity. Certainty of evidence was very low.

**Conclusions:** In pooled US observational data, rural patients receiving emergency and trauma care had about 23% higher odds of short-term death than urban patients, most consistently among injured children. The evidence is inconsistent, rests on a single-database search with several reports assessed from abstracts only, and should be treated as preliminary. Why rural patients die more often, and at what point in the pathway from injury to definitive care, remains unanswered.

---

For many rural Americans, the emergency department (ED) is the only door into the health system that is always open. Rural populations rely heavily on emergency care, and that reliance has grown over time.^1^ The hospitals behind those doors are not equipped alike. A systematic review of rural EDs in developed countries found wide variation in staffing, physical resources, diagnostic access, and specialist services, all of which limit the capacity to treat high-acuity patients.^2^

It would be reasonable to expect these differences to cost lives. The published evidence does not settle the matter. National analyses have reported higher mortality for rural residents after injury, similar mortality after rural and urban ED visits, and lower early mortality in rural hospitals. These studies differ in patient population, clinical condition, the way rurality is defined, and the time point at which death is counted, so no single study can show whether rural emergency care is consistently associated with higher mortality, or by how much.

The goal of this study was to determine whether mortality differs between rural and urban emergency and trauma care settings in the United States and to estimate the size of any difference. Among patients receiving emergency or trauma care in the United States, we asked whether care in, or residence in, a rural setting is associated with a difference in short-term mortality compared with an urban setting.

## METHODS

### Protocol and reporting

This review is reported in line with the PRISMA 2020 statement.^3^ The review was not registered, and no protocol was published. An earlier version included four studies and used narrative synthesis only; the present version replaces it with a docu-mented database search and a meta-analysis.

### Eligibility criteria

We included observational studies of patients receiving ED, emergency surgical, or acute trauma care in the United States that compared rural with urban settings and reported short-term mortality (ED, in-hospital, 48-hour, or 30-day) for individual patients. Rurality could be defined by hospital location, patient residence, or region, including multilevel scales. Studies had to be in English and published in 2010 or later; dissertations were eligible. We excluded studies conducted outside the United States, general inpatient cohorts not defined by emergency care, county-level population death rates, studies without a rural-urban comparison or an extractable mortality estimate, reviews, editorials, and case reports.

### Search strategy

PubMed was searched on October 11, 2026, with the following string:

~~~
(rural[tiab] AND urban[tiab]) AND (mortality[tiab] OR
death[tiab]) AND (“emergency department”[tiab] OR “emergency
departments”[tiab] OR “Emergency Service, Hospital”[MeSH] OR
“trauma center”[tiab] OR “trauma centers”[tiab] OR “Wounds and
Injuries”[MeSH]) AND “United States”[MeSH] AND 2010:2026[dp]
~~~

Additional records were identified through nonsystematic web searching and from the earlier version of this review. No other bibliographic database was searched.

### Study selection

Records from PubMed were screened by title because the export did not include abstracts. Records that compared rural and urban patients or hospitals and could plausibly report patient-level mortality after emergency or trauma care were sought for full assessment. Reports were assessed in full text where it was available to the authors and from the published abstract otherwise. A large language model (LLM) was used to assist with title screening, eligibil-ity assessment, and data extraction. One author (AE) screened the records and verified every LLM-assisted decision and all extracted data against the source reports. Screening was not performed independently in duplicate.

### Data extraction

From each included study, we extracted the first author, year, data source, study period, population, sample size, definition of rurality, mortality outcome, crude mortality by setting where reported, and the most fully adjusted OR for rural vs urban mortality with its 95% CI. Where a study used a multilevel rurality scale, we extracted the comparison of the most rural category with a reported estimate against the most urban category. Where urban hospitals were divided by teaching status, we extracted the comparison of rural with urban nonteaching hospitals as the closer comparator.

### Risk of bias and certainty of evidence

All included studies were observational. A validated item-level risk-of-bias instrument was not applied because several studies were available only as abstracts. Instead, for each study, we recorded three features most likely to bias a rural-urban mortality comparison: whether the estimate was adjusted for injury or illness severity, how interhospital transfers were handled, and whether rurality described the patient or the hospital. Certainty of evidence for the primary outcome was rated with the GRADE approach, which starts observational evidence at low certainty and rates down for risk of bias, inconsistency, indirectness, imprecision, and publication bias.

### Statistical analysis

The effect measure was the OR for death in rural compared with urban settings. Log ORs and standard errors were derived from the reported 95% CIs and pooled with a DerSimonian-Laird random-effects model.^4^ Heterogeneity was summarized with the Cochran Q, I^2^, and *τ*^2^, and a 95% prediction interval was calculated from the t distribution with k − 2 degrees of freedom. The primary analysis pooled studies of injury and trauma; a secondary analysis added studies of other emergency conditions. Two pooled estimates were not adjusted by regression: one study reported matched mortality proportions without an overall OR in the main text, and an OR was calculated from the reported percentages and group sizes; another reported an unadjusted OR. Studies that reported no CI were described narratively and not pooled.

Because few studies were pooled and heterogeneity was expected, CIs were also calculated with the Hartung-Knapp method, which is more conservative.^5^ Further sensitivity analyses excluded the two unadjusted estimates, removed one trauma study at a time, and restricted the analysis to studies assessed in full text. Three exploratory sub-group analyses, chosen after data extraction, compared studies by rurality definition (patient residence or region vs hospital location), age group (pediatric vs adult or all ages, trauma studies only), and geographic scope (national vs state or regional). Subgroup differences were tested with a z test on the pooled log ORs. Publication bias was not formally assessed because heterogeneity was too high for funnel plot asymmetry to be interpretable. Certainty of evidence followed GRADE.^6^ Analyses were performed in Python 3.

## RESULTS

### Study selection

The PubMed search returned 155 records, and six further records came from web searching and the earlier version of this review (Fig 1). Title screening excluded 134 PubMed records, most of them county-level death rate reports or studies with no rural-urban comparison. All 21 remaining reports were assessed, nine in full text and 12 from the abstract. Ten were excluded: eight made no rural-urban comparison of patient mortality, and two reported no adjusted or overall estimate. Of the six records from other sources, three were excluded. Excluded reports and reasons are listed in Supplementary Table I (online only). Fourteen studies were included in the review and 12 in the meta-analysis.

**Fig 1.**
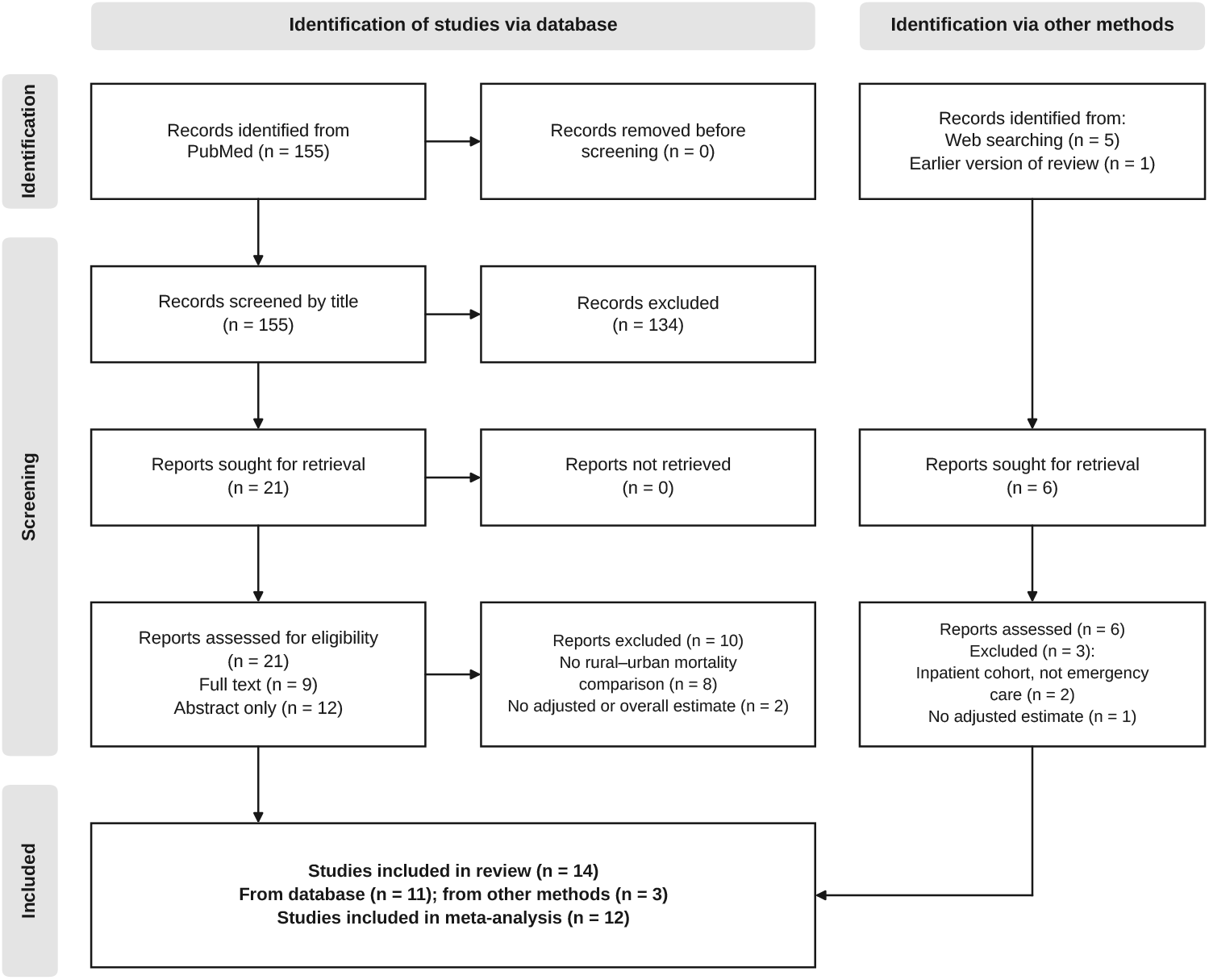
PRISMA 2020 flow diagram of study selection.

### Study characteristics

The 14 included studies^7-20^ were published between 2011 and 2026 (Table I). Eight examined injury or trauma, and six examined other emergency populations: ruptured abdominal aortic aneurysm (two studies), emergency general surgery admissions, Medicare ED visits for any diagnosis (two studies), and cardiovascular ED visits. Eleven used national administrative data, two used state data, and one was a prospective regional cohort. Rurality was defined by patient residence or region in seven studies and by hospital location in seven. Four included studies were assessed from the abstract only,^7,12,18,20^ and one is a doctoral dissertation that has not been peer reviewed.^20^

Eight of the 14 studies adjusted for a measure of injury or illness severity (Table II). Handling of interhospital transfers varied: three studies excluded transferred patients, two followed patients across facilities or attributed outcomes to the first ED, and the others did not capture deaths after transfer or did not report their approach.

### Mortality after injury and trauma

Seven injury and trauma studies were pooled (Fig 2). Rural setting was associated with higher odds of death (OR, 1.27; 95% CI, 1.07-1.52). Heterogeneity was substantial (Q = 38.3; I^2^ = 84%; *τ*^2^ = 0.036), and the 95% prediction interval was 0.74 to 2.19. Five studies found significantly higher odds of death for rural patients,^9,10,16–18^ and two found no significant difference.^11,15^

**Fig 2.**
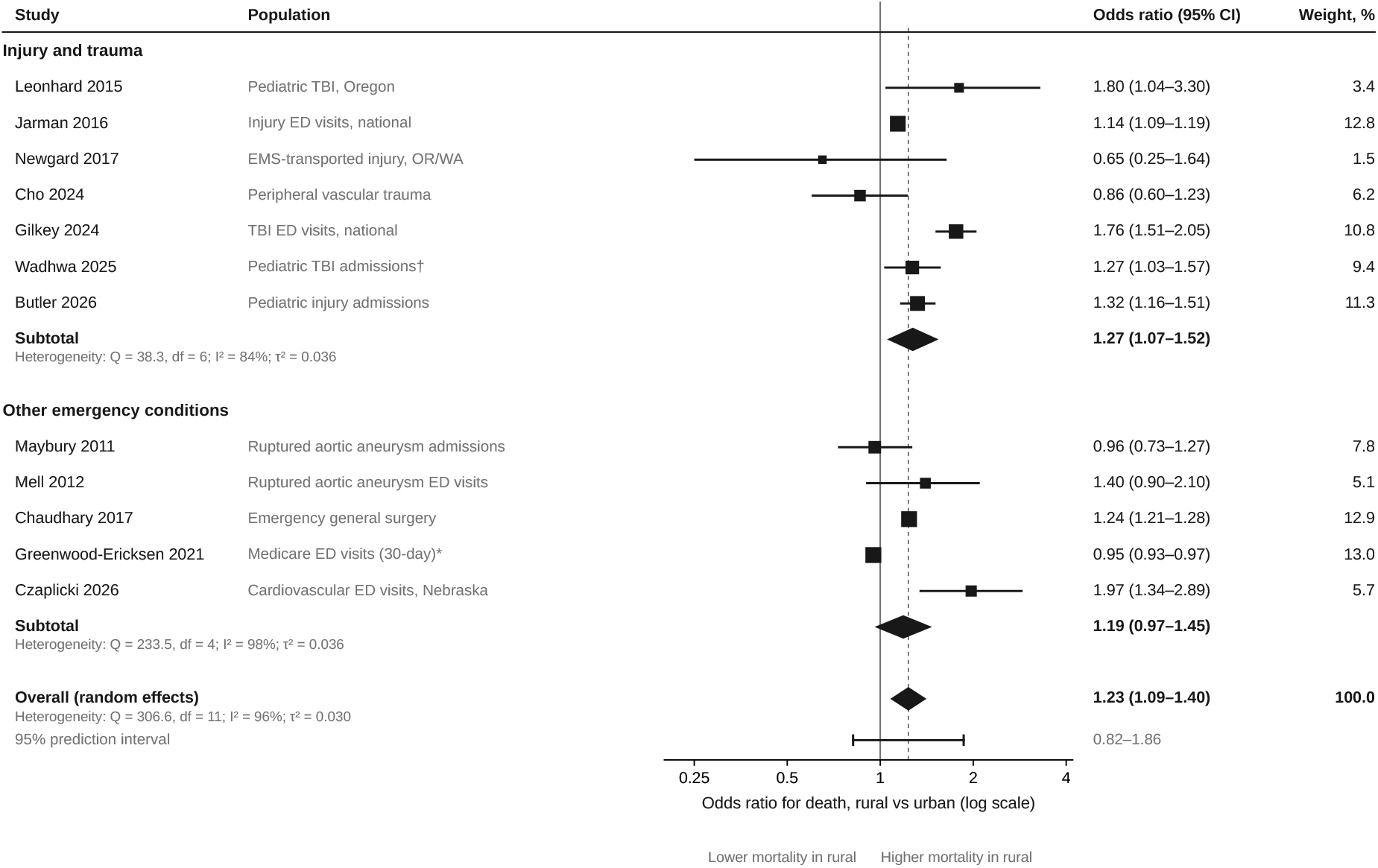
Random-effects meta-analysis of the odds of death in rural vs urban emergency and trauma care settings. Squares are sized by weight in the overall analysis, and horizontal lines show 95% confidence intervals (CIs). Diamonds show pooled estimates. The dashed line marks the overall pooled estimate, and the bar beneath the overall diamond shows the 95% prediction interval. ^*^Odds ratio calculated from reported percentages. †Unadjusted odds ratio. *ED*, Emergency department; *EMS*, emergency medical services; *TBI*, traumatic brain injury.

In the prospective cohort of Newgard et al, overall mortality did not differ significantly between rural and urban regions (1.44% vs 0.89%; P = .09), but 89.6% of rural deaths occurred within 24 hours compared with 64% of urban deaths.^11^ In the study of Cho et al, in-hospital mortality was similar (2.5% vs 2.6%), whereas rural patients had higher odds of amputation (adjusted OR, 1.85; 95% CI, 1.47-2.32).^15^ One further trauma study could not be pooled. Gilbert Henderson reported lower 48-hour mortality in rural than in urban hospitals (0.8% vs 1.4%; adjusted OR, 0.70) among admissions that excluded all transferred patients.^20^

### Other emergency conditions

Five studies of other emergency populations were pooled, and their results conflicted. Rural hospitals had higher in-hospital mortality for emergency general surgery (OR, 1.24; 95% CI, 1.21-1.28),^12^ and rural Nebraska counties had higher in-hospital mortality after cardiovascular ED visits (OR, 1.97; 95% CI, 1.34-2.89).^19^ For ruptured abdominal aortic aneurysm, death in the ED was more frequent in rural hospitals (12.7% vs 7.0% in urban nonteaching hospitals), although the adjusted difference was not significant (OR, 1.40; 95% CI, 0.90-2.10),^8^ and in-hospital mortality among admitted patients was similar (OR, 0.96; 95% CI, 0.73-1.27).^7^ The pooled estimate for the five studies was not significant (OR, 1.19; 95% CI, 0.97-1.45; I^2^ = 98%).

The two studies of Medicare ED visits disagreed. A matched cohort found slightly lower 30-day mortality after rural than after urban visits (3.9% vs 4.1%).^14^ In that study, rural patients were transferred more often (6.2% vs 2.0%), mortality was similar for potentially life-threatening diagnoses, and mortality was higher after rural visits for symptom-based diagnoses such as chest pain (OR, 1.54; 95% CI, 1.25-1.89). In contrast, Burke et al reported higher adjusted 30-day mortality after visits to rural than to urban EDs in both 2009 (6.2% vs 5.6%) and 2016 (5.0% vs 4.2%), with no CIs for the difference, so the study was not pooled.^13^ The two cohorts overlap in source and years and differ in method: one matched visits on diagnosis and patient characteristics, and the other used regression adjustment across all visits.

### Overall and sensitivity analyses

Across all 12 studies, the pooled OR was 1.23 (95% CI, 1.09-1.40; Q = 306.6; I^2^ = 96%), with a 95% prediction interval of 0.82 to 1.86. Eight of the 12 estimates were above 1.00, and seven of those were statistically significant. Sensitivity and subgroup analyses are shown in Table III.

The overall result was robust to the analytic method, and the trauma result was borderline. With Hartung-Knapp intervals, the overall estimate remained significant (95% CI, 1.05-1.45), and the primary trauma estimate narrowly included no difference (95% CI, 0.99-1.64). In the trauma analysis, removing one study at a time gave pooled ORs between 1.19 and 1.34 (Fig 3), and the lower confidence limit fell below 1.00 only when the study of Butler et al was removed (95% CI, 0.99-1.59). Removing the study of Gilkey et al reduced heterogeneity the most (I^2^ = 56%).

**Fig 3.**
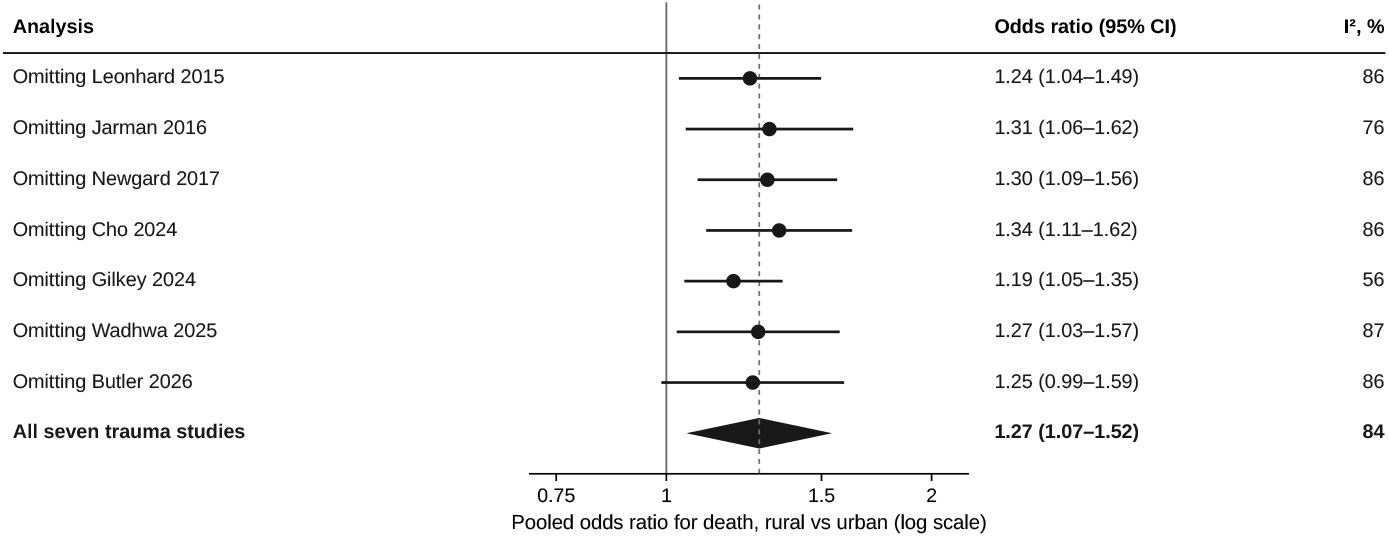
Leave-one-out sensitivity analysis of the primary injury and trauma meta-analysis. Each row shows the random-effects pooled odds ratio and I^2^ after the named study is omitted. The dashed line marks the estimate from all seven studies. *CI*, Confidence interval.

None of the subgroup differences was statistically significant. Pooled estimates were similar whether rurality was defined by the patient or by the hospital (1.25 vs 1.22). The three pediatric trauma studies agreed closely (OR, 1.32; 95% CI, 1.18-1.47; I^2^ = 0%) and remained significant under the Hartung-Knapp method. Two of them drew on national inpatient data from overlapping years,^17,18^ so they are not fully independent.

### Certainty of evidence

Certainty that rural setting is associated with higher short-term mortality was rated very low. Observational evidence starts at low certainty, and we rated down for inconsistency (I^2^ above 80% and a prediction interval that includes no difference) and for risk of bias (variable handling of transfers, unvalidated appraisal, and assessment of several reports from abstracts only).

## DISCUSSION

In this meta-analysis of 12 US observational studies, rural patients receiving emergency and trauma care had about 23% higher odds of short-term death than urban patients. Eight of the 12 estimates were above 1.00, and seven of those were statistically significant. The overall result held under the more conservative Hartung-Knapp method and when the two unadjusted estimates were removed. The studies nevertheless disagreed considerably: individual estimates ranged from 0.65 to 1.97, I^2^ exceeded 80% in the main analyses, and the prediction intervals included no difference. The average effect therefore should not be read as the effect expected in any particular setting, and certainty in it is very low.

The most consistent signal was in children. Three pediatric trauma studies found higher mortality for rural children with almost no heterogeneity,^9,17,18^ in line with the limited availability of pediatric trauma care outside large urban areas. This finding should be read with caution because two of the three studies drew on overlapping national data and one estimate was unadjusted. The gradient reported by Butler et al, in which odds of death rose stepwise from large urban to small urban, micropolitan, and rural counties, suggests that a binary rural-urban split hides meaningful variation.^18^

We expected the definition of rurality to explain part of the disagreement. Studies that classify patients by where they live capture the whole pathway, including injury mechanism, prehospital time, and distance to a trauma center, whereas studies that classify hospitals capture care at one facility. The data did not support this expectation; pooled estimates were similar for the two definitions.

Timing and place of death may matter more. Newgard et al found that most rural trauma deaths occurred within 24 hours.^11^ Mell et al found that patients with a ruptured aortic aneurysm were more likely to die in the ED at rural hospitals,^8^ whereas Maybury et al found similar mortality among those who survived to admission.^7^ Together, these observations point toward early, time-sensitive care as the period of greatest rural risk.

**Table I.** Characteristics and findings of included studies.

| Study | Data source and years | Population (No.) | Rurality defined by | Mortality outcome | OR (95% CI) | Main concern for bias |
| --- | --- | --- | --- | --- | --- | --- |
| Maybury 2011<sup>7</sup> | Nationwide Inpatient Sample, 2001-2007 | Admissions with ruptured abdominal aortic aneurysm (not stated in abstract) | Hospital | In-hospital death | 0.96 (0.73-1.27) | Abstract only; transferred patients excluded |
| Mell 2012<sup>8</sup> | Nationwide Emergency Department Sample, 2006-2008 | ED visits for ruptured abdominal aortic aneurysm (18,363) | Hospital | Death in the ED | 1.40 (0.90-2.10) | Reference group was urban nonteaching hospitals; not adjusted for severity |
| Leonhard 2015<sup>9</sup> | Oregon Trauma Registry, 2009-2012 | Children with traumatic brain injury (2794) | Residence | Death recorded in registry | 1.80 (1.04-3.30) | Single state; 76 deaths |
| Jarman 2016<sup>10</sup> | Nationwide Emergency Department Sample, 2009-2010 | Injury ED visits (8,673,213) | Residence | ED or in-hospital death | 1.14 (1.09-1.19) | Transferred patients excluded |
| Newgard 2017<sup>11</sup> | Prospective EMS cohort, Oregon and Washington, 2011 | Injured patients served by 9-1-1 EMS (67,047 enrolled; outcomes from a probability sample of 17,633) | Region | In-hospital death | 0.65 (0.25-1.64) | Two rural counties; 29 rural deaths |
| Chaudhary 2017<sup>12</sup> | National Inpatient Sample, 2007-2011 | Emergency general surgery admissions (3,749,265) | Hospital | In-hospital death | 1.24 (1.21-1.28) | Abstract only |
| Burke 2020<sup>13</sup> | Medicare sample, 2009-2016 | ED visits by beneficiaries aged 65 years or older (15,416,385) | Hospital | 30-day all-cause death | Not reported | Adjusted rates without CIs; not pooled |
| Greenwood-Ericksen 2021<sup>14</sup> | 20% Medicare sample, 2011-2015 | Matched ED visits (473,152 rural; 473,152 urban) | Hospital | 30-day all-cause death | 0.95 (0.93-0.97)<sup>a</sup> | OR derived from rounded percentages |
| Cho 2024<sup>15</sup> | Nationwide Readmissions Database, 2016-2020 | Adults treated for peripheral vascular trauma (29,083) | Residence | In-hospital death | 0.86 (0.60-1.23) | Operative patients only |
| Gilkey 2024<sup>16</sup> | Nationwide Emergency Department Sample, 2016-2020 | Traumatic brain injury ED visits (about 500,000 per year) | Hospital | ED or in-hospital death | 1.76 (1.51-2.05) | Reference group was urban nonteaching hospitals |
| Wadhwa 2025<sup>17</sup> | National Inpatient Sample, 2011-2020 | Children admitted with traumatic brain injury (45,109) | Residence | In-hospital death | 1.27 (1.03-1.57)<sup>b</sup> | Unadjusted; most rural category not reported |
| Butler 2026<sup>18</sup> | Kids' Inpatient Database, 2016, 2019, 2022 | Injured children admitted to hospital (402,255 weighted) | Residence | Inpatient death | 1.32 (1.16-1.51) | Abstract only |
| Czaplicki 2026<sup>19</sup> | Nebraska state ED and inpatient databases, 2016-2020 | Cardiovascular ED visits (47,369; 25,190 admitted) | County | In-hospital death among admitted | 1.97 (1.34-2.89) | Single state; deaths after transfer not captured |
| Gilbert Henderson 2026<sup>20</sup> | National Inpatient Sample, 2022 | Trauma admissions (182,560) | Hospital | 48-hour in-hospital death | 0.70 (CI not reported) | Transfers excluded; dissertation; not pooled |

CI, Confidence interval; ED, emergency department; EMS, emergency medical services; OR, odds ratio for death, rural vs urban. <sup>a</sup>Calculated from the reported 30-day mortality of 3.9% and 4.1%; not adjusted beyond the propensity matching. <sup>b</sup>Unadjusted; micropolitan counties compared with central counties of metropolitan areas of at least 1 million.

**Table II.** Features of included studies that affect a rural-urban mortality comparison.

| Study | Adjusted for severity | Other adjustment | Handling of transfers | Source assessed |
| --- | --- | --- | --- | --- |
| Maybury 2011<sup>7</sup> | Not stated in abstract | Patient, hospital, and health system variables | Patients transferred in were excluded | Abstract |
| Mell 2012<sup>8</sup> | No | Age, sex, comorbidity, insurance, region, ED volume, trauma designation | Deaths after transfer not captured | Full text |
| Leonhard 2015<sup>9</sup> | Yes (Injury Severity Score, injury type) | Age, sex, race, insurance | Not reported | Full text |
| Jarman 2016<sup>10</sup> | Yes (Injury Severity Score) | Age, sex, comorbidity, injury type, trauma center level, region | Transferred patients excluded | Full text |
| Newgard 2017<sup>11</sup> | Yes (trauma risk-adjustment model) | Standard trauma confounders | Followed across hospitals | Full text |
| Chaudhary 2017<sup>12</sup> | Not stated in abstract | Not stated in abstract | Not stated in abstract | Abstract |
| Burke 2020<sup>13</sup> | Yes (diagnosis-based severity of illness) | Age, sex, race, Medicaid enrollment, chronic conditions, hospital characteristics | Not reported | Full text |
| Greenwood-Ericksen 2021<sup>14</sup> | No direct severity measure | Matched on demographics, prior ED use, comorbidity, diagnosis | Outcomes attributed to the first ED | Full text |
| Cho 2024<sup>15</sup> | Yes (Trauma Mortality Prediction Model) | Age, sex, insurance, comorbidity, injury type and mechanism, hospital factors | Transfers included (2.8% rural, 2.5% urban) | Full text |
| Gilkey 2024<sup>16</sup> | Yes (Injury Severity Score) | Age, sex, hospital type, trauma center level, study period | Deaths after transfer not captured | Full text |
| Wadhwa 2025<sup>17</sup> | No | None | Not reported | Full text |
| Butler 2026<sup>18</sup> | Yes (risk of mortality) | Survey design | Not stated in abstract | Abstract |
| Czaplicki 2026<sup>19</sup> | Not confirmed | Multivariable model | Deaths after transfer not captured | Full text |
| Gilbert Henderson 2026<sup>20</sup> | Injury complexity | Patient and hospital covariates | Transferred patients excluded | Abstract |

ED, Emergency department.

**Table III.** Sensitivity and exploratory subgroup analyses.

| Analysis | Studies, No. | OR (95% CI) | Hartung-Knapp 95% CI | $I^2$, % | P for subgroup difference |
| --- | --- | --- | --- | --- | --- |
| All studies | 12 | 1.23 (1.09-1.40) | 1.05-1.45 | 96 |  |
| Injury and trauma (primary) | 7 | 1.27 (1.07-1.52) | 0.99-1.64 | 84 |  |
| Other emergency conditions | 5 | 1.19 (0.97-1.45) | 0.84-1.67 | 98 |  |
| Excluding the two unadjusted estimates<sup>14,17</sup> | 10 | 1.28 (1.16-1.41) | 1.08-1.52 | 82 |  |
| Full-text studies only | 9 | 1.26 (1.08-1.47) | 0.99-1.59 | 94 |  |
| Trauma, excluding Gilkey<sup>16</sup> | 6 | 1.19 (1.05-1.35) | 0.98-1.44 | 56 |  |
| Rurality by patient residence or region | 7 | 1.25 (1.08-1.45) | 0.99-1.58 | 68 | .84 |
| Rurality by hospital location | 5 | 1.22 (1.00-1.48) | 0.87-1.69 | 99 |  |
| Trauma, pediatric | 3 | 1.32 (1.18-1.47) | 1.09-1.60 | 0 | .47 |
| Trauma, adult or all ages | 4 | 1.16 (0.84-1.62) | 0.64-2.13 | 91 |  |
| National data | 9 | 1.19 (1.05-1.36) | 1.01-1.41 | 97 | .37 |
| State or regional data | 3 | 1.52 (0.90-2.57) | 0.41-5.68 | 57 |  |

CI, Confidence interval; OR, random-effects pooled odds ratio for death, rural vs urban.

Handling of transfers is a likely source of disagreement. Rural hospitals transfer many of their sickest patients: transfer rates were about three times higher after rural than after urban ED visits in one cohort,^14^ and rural hospitals accounted for half of all transfers for ruptured aortic aneurysm in another.^8^ When transferred patients are excluded, severely ill rural patients drop out of the rural group, which can make rural hospitals appear safer than they are. The lower 48-hour mortality reported by Gilbert Henderson^20^ more likely reflects this case-mix effect than a protective effect of rural care.

The two Medicare studies illustrate how much method matters. Using overlapping data, a diagnosis-matched comparison found slightly lower mortality after rural ED visits,^14^ whereas a regression-adjusted comparison across all visits found higher mortality.^13^ The pooled anal-ysis includes only the first of these because the second reported no CIs, so the overall estimate may understate the rural disadvantage in older adults.

This review establishes that a mortality gap is likely but cannot say where in the pathway it arises. The included studies measured death after patients reached a hospital, and none was designed to separate what happens in the hospital from what happens before arrival. The one study that followed patients from the scene suggests that the earlier steps deserve attention. In the cohort of Newgard et al, field triage identified 65.2% of rural patients who went on to need early critical resources, compared with 80.5% of urban patients, and only 29.4% of those rural patients were taken first to a major trauma center, compared with 88.7% in urban regions.^11^ Whether the rural excess in deaths is driven by the recognition of severe injury in the field, the choice of first hospital, or the delay to definitive care is the central unanswered question, and it is one that administrative hospital data cannot resolve. Studies that follow rural patients from the point of injury, with outcomes attributed to the first triage decision, are the logical next step.

These findings support directing attention to access to time-critical specialist care and not to hospital location as such. Regional trauma systems, emergency medical services, field triage, interfacility transfer, and telemedicine support are plausible targets, particularly for injured children in rural counties. This review did not evaluate any of these interventions, so these are directions for research and not evidence-based recommendations. Future studies should also report a common rurality classification, a common mortality end point, and outcomes for transferred patients attributed to the first treating hospital.

This review has important limitations, and its results should be treated as preliminary. Only one bibliographic database was searched, records were screened by title, and additional studies were found by nonsystematic web searching, so eligible studies have probably been missed. Twelve of the 21 database reports were assessed from the abstract, including four of the 14 included studies, and details of adjustment and transfer handling could not be confirmed for these. The review was not registered, and screening was performed by a single author with LLM assistance and not independently in duplicate. No validated item-level risk-of-bias instrument was applied, and the GRADE rating was made by the review team without independent duplication. Populations, rurality definitions, comparator groups, and mortality end points differed, the pooled estimates combine them, and the subgroup analyses were exploratory and underpowered. Most studies drew on Healthcare Cost and Utilization Project databases or Medicare claims, so some patients contribute to more than one estimate and the studies are not fully independent. Two pooled ORs were not regression adjusted, one of them calculated from rounded percentages. Finally, all studies were observational, so residual confounding by injury severity, comorbidity, and transfer is likely, and no causal conclusion can be drawn.

## CONCLUSIONS

US observational studies suggest that rural patients receiving emergency and trauma care have higher odds of short-term death than urban patients, by about 23% on average, with the most consistent evidence in injured children. The evidence is inconsistent across settings, and certainty is very low. A registered review with a multi-database search, duplicate screening, and formal risk-of-bias assessment is needed to confirm the size of the difference, and studies that follow patients from the scene of injury are needed to explain it.

## Supporting information

Supplementary Tables I and II: Excluded reports and PRISMA 2020 checklist

## Data Availability

All data analyzed in this study were extracted from previously published studies cited in the manuscript and are contained in the manuscript (Tables I–III) and its supplementary material. The PubMed search export and analysis code are available upon reasonable request to the corresponding author.

## REFERENCES

1. Greenwood-Ericksen MB, Kocher K. Trends in emergency department use by rural and urban populations in the United States. JAMA Netw Open 2019;2:e191919.

2. Kerr L, Kealy B, Lim D, Walters L. Rural emergency departments: a systematic review to develop a resource typology relevant to developed countries. Aust J Rural Health 2021;29:7–20.

3. Page MJ, McKenzie JE, Bossuyt PM, Boutron I, Hoffmann TC, Mulrow CD, et al. The PRISMA 2020 statement: an updated guideline for reporting systematic reviews. BMJ 2021;372:n71.

4. DerSimonian R, Laird N. Meta-analysis in clinical trials. Control Clin Trials 1986;7:177–88.

5. IntHout J, Ioannidis JP, Borm GF. The Hartung-Knapp-Sidik-Jonkman method for random effects meta-analysis is straightforward and considerably outperforms the standard DerSimonian-Laird method. BMC Med Res Methodol 2014;14:25.

6. Guyatt GH, Oxman AD, Vist GE, Kunz R, Falck-Ytter Y, Alonso-Coello P, et al. GRADE: an emerging consensus on rating quality of evidence and strength of recommendations. BMJ 2008;336:924–6.

7. Maybury RS, Chang DC, Freischlag JA. Rural hospitals face a higher burden of ruptured abdominal aortic aneurysm and are more likely to transfer patients for emergent repair. J Am Coll Surg 2011;212:1061–7.

8. Mell MW, Callcut RA, Bech F, Delgado MK, Staudenmayer K, Spain DA, et al. Predictors of emergency department death for patients presenting with ruptured abdominal aortic aneurysms. J Vasc Surg 2012;56:651–5.

9. Leonhard MJ, Wright DA, Fu R, Lehrfeld DP, Carlson KF. Urban/rural disparities in Oregon pediatric traumatic brain injury. Inj Epidemiol 2015;2:32.

10. Jarman MP, Castillo RC, Carlini AR, Kodadek LM, Haider AH. Rural risk: geographic disparities in trauma mortality. Surgery 2016;160:1551–9.

11. Newgard CD, Fu R, Bulger E, Hedges JR, Mann NC, Wright DA, et al. Evaluation of rural vs urban trauma patients served by 9-1-1 emergency medical services. JAMA Surg 2017;152:11–8.

12. Chaudhary MA, Shah AA, Zogg CK, Changoor N, Chao G, Nitzschke S, et al. Differences in rural and urban outcomes: a national inspection of emergency general surgery patients. J Surg Res 2017;218:277–84.

13. Burke LG, Epstein SK, Burke RC, Orav EJ, Jha AK. Trends in mortality for Medicare beneficiaries treated in the emergency department from 2009 to 2016. JAMA Intern Med 2020;180:80–8.

14. Greenwood-Ericksen M, Kamdar N, Lin P, George N, Myaskovsky L, Crandall C, et al. Association of rural and critical access hospital status with patient outcomes after emergency department visits among Medicare beneficiaries. JAMA Netw Open 2021;4:e2134980.

15. Cho NY, Vadlakonda A, Curry J, Tran Z, Tillou A, de Virgilio C, et al. Association of rurality with short-term outcomes of peripheral vascular trauma. Surgery 2024;176:205–10.

16. Gilkey S, Armstrong M, Alexander R, Osong B, Sribnick EA, Stanley RM, et al. Impact of COVID-19 pandemic on traumatic brain injury emergency department visits, interfacility transfer and mortality in the United States, 2016-2020: a cross-sectional study. BMJ Open 2024;14:e090141.

17. Wadhwa A, Purohit S, Ramirez-Velandia F, Enriquez-Marulanda A, Ogilvy CS. Socioeconomic disparities in the clinical outcomes of pediatric traumatic brain injury: a nationwide analysis of the past decade. World Neurosurg 2025;194:123420.

18. Butler EK, Johnson R, Saltzman DA, Segura BJ. Beyond rural versus urban: a gradient of mortality risk among injured children across a four-level rurality scale. J Trauma Acute Care Surg 2026;101:475–82.

19. Czaplicki AM, Dilsaver DB, Kim SY, Kim MH. Differential patterns of urban and rural cardiovascular emergency department utilization and clinical outcomes. Front Disaster Emerg Med 2026;4:1766652.

20. Gilbert Henderson K. Analyzing variations in 48-hour mortality rates following hospital admission for trauma injuries across urban and rural hospitals [dissertation]. Charleston, SC: Medical University of South Carolina; 2026.

