## Supplementary Tables I and II: Excluded reports and PRISMA 2020 checklist for "Mortality in Rural and Urban Emergency and Trauma Care in the United States: A Systematic Review and Meta-Analysis"

**Supplementary Table I (online only).** Reports assessed for eligibility and excluded, with reasons

| *Report* | *Source* | *Assessed from* | *Reason for exclusion* |
|---|---|---|---|
| Byrd 2025[1] | PubMed | Full text | No rural-urban mortality estimate |
| de Jager 2020[2] | PubMed | Abstract | Income effects reported within rural and urban strata; no rural-urban comparison |
| Kallies 2025[3] | PubMed | Abstract | Mortality estimate reported for one rural subgroup only |
| Kallies 2026[4] | PubMed | Abstract | Crude mortality by six area types; no adjusted or overall rural-urban estimate |
| Ortega 2013[5] | PubMed | Abstract | Describes deaths only; no denominators for a mortality comparison |
| Harwell 2019[6] | PubMed | Abstract | Compares rural laparotomy with transfer; no urban comparator |
| Patel 2021[7] | PubMed | Abstract | No rural-urban mortality comparison |
| Anderson 2014[8] | PubMed | Abstract | County-level injury rates; no mortality comparison by rurality |
| Bir 2015[9] | PubMed | Abstract | Hospital location analyzed for discharge outcome, not mortality |
| Wild 2017[10] | PubMed | Abstract | Compares direct admission with transfer; no urban comparator |
| Chang 2023[11] | Web search | Full text | Inpatient sepsis cohort, not defined by emergency care |
| Loccoh 2022[12] | Web search | Abstract | Inpatient cardiovascular cohort reporting hazard ratios |
| Marek 2018[13] | Web search | Full text | Two-center comparison with no adjusted estimate |

**Supplementary Table II (online only).** PRISMA 2020 checklist

| *Item* | *Topic* | *Where reported* |
|---|---|---|
| 1 | Title | Title |
| 2 | Abstract | Abstract |
| 3 | Rationale | Introduction |
| 4 | Objectives | Introduction, final paragraph |
| 5 | Eligibility criteria | Methods, Eligibility criteria |
| 6 | Information sources | Methods, Search strategy |
| 7 | Search strategy | Methods, Search strategy (full PubMed string) |
| 8 | Selection process | Methods, Study selection |
| 9 | Data collection process | Methods, Study selection; Data extraction |
| 10a | Data items: outcomes | Methods, Data extraction |
| 10b | Data items: other variables | Methods, Data extraction |
| 11 | Study risk of bias assessment | Methods, Risk of bias and certainty of evidence (no validated instrument applied) |
| 12 | Effect measures | Methods, Statistical analysis |
| 13a | Eligibility for each synthesis | Methods, Statistical analysis |
| 13b | Data preparation | Methods, Statistical analysis (derived odds ratio) |
| 13c | Tabulation and display | Tables I to III; Figs 2 and 3 |
| 13d | Synthesis methods | Methods, Statistical analysis |
| 13e | Exploring heterogeneity | Methods, Statistical analysis (subgroup analyses) |
| 13f | Sensitivity analyses | Methods, Statistical analysis |
| 14 | Reporting bias assessment | Not assessed; stated in Methods |
| 15 | Certainty assessment | Methods, Risk of bias and certainty of evidence |
| 16a | Study selection | Results, Study selection; Fig 1 |
| 16b | Excluded studies | Supplementary Table I |
| 17 | Study characteristics | Table I |
| 18 | Risk of bias in studies | Table II |
| 19 | Results of individual studies | Table I; Fig 2 |
| 20a | Characteristics of syntheses | Results, Mortality after injury and trauma; Other emergency conditions |
| 20b | Results of syntheses | Fig 2; Table III |
| 20c | Causes of heterogeneity | Table III |
| 20d | Sensitivity analyses | Table III; Fig 3 |
| 21 | Reporting biases | Not assessed |
| 22 | Certainty of evidence | Results, Certainty of evidence |
| 23a | Interpretation | Discussion |
| 23b | Limitations of evidence | Discussion, final paragraph |
| 23c | Limitations of review processes | Discussion, final paragraph |
| 23d | Implications | Discussion |
| 24a | Registration | Methods, Protocol and reporting (not registered) |
| 24b | Protocol | Methods, Protocol and reporting (no protocol) |
| 24c | Amendments | Not applicable |
| 25 | Support | No funding; declared at submission |
| 26 | Competing interests | None; declared at submission |
| 27 | Availability of data, code, and materials | Tables I and II; Supplementary Table I |

### REFERENCES


1. Byrd T, Kass N, Lu L, Silver D, Boland S, Brown JB. Association of injury-related mortality with unique domains of social determinants of health across the urban-rural continuum. Surgery 2025;187:109607.
2. de Jager E, Chaudhary MA, Rahim F, Jarman MP, Uribe-Leitz T, Havens JM, et al. The impact of income on emergency general surgery outcomes in urban and rural areas. J Surg Res 2020;245:629-35.
3. Kallies KJ, Koehne WJ, Tomas CW, Tarima S, Beckman MA, Beyer KM, et al. A comparison of traumatic injury characteristics in rural and urban adults living in Wisconsin. J Rural Health 2025;41:e12922.
4. Kallies KJ, Beyer KM, Koehne WJ, Tomas CW, Tarima S, Beckman MA, et al. Pediatric traumatic injury in rural and urban areas of Wisconsin. Pediatr Emerg Care 2026;42:571-6.
5. Ortega HW, Vander Velden H, Krause E, Reid S. Traumatic deaths in children: is there a difference between urban and rural populations? Pediatr Emerg Care 2013;29:36-8.
6. Harwell PA, Reyes J, Helmer SD, Haan JM. Outcomes of rural trauma patients who undergo damage control laparotomy. Am J Surg 2019;218:490-5.
7. Patel SJ, Badolato GM, Parikh K, Iqbal SF, Goyal MK. Sociodemographic factors and outcomes by intent of firearm injury. Pediatrics 2021;147:e2020011957.
8. Anderson BL, Pomerantz WJ, Gittelman MA. Intentional injuries in young Ohio children: is there urban/rural variation? J Trauma Acute Care Surg 2014;77(3 Suppl 1):S36-40.
9. Bir SC, Maiti TK, Ambekar S, Nanda A. Incidence, hospital costs and in-hospital mortality rates of epidural hematoma in the United States. Clin Neurol Neurosurg 2015;138:99-103.
10. Wild J, Younus JM, Malekpour M, Neuhaus N, Widom K, Rapp M, et al. The effect of interhospital transfers on the outcome of rural trauma. Am Surg 2017;83:39-44.
11. Chang J, Medina M, Kim SJ. Is patients’ rurality associated with in-hospital sepsis death in US hospitals? Front Public Health 2023;11:1169209.
12. Loccoh EC, Joynt Maddox KE, Wang Y, Kazi DS, Yeh RW, Wadhera RK. Rural-urban disparities in outcomes of myocardial infarction, heart failure, and stroke in the United States. J Am Coll Cardiol 2022;79:267-79.
13. Marek AP, Nygaard RM, Cohen EM, Polites SF, Sirany AE, Wildenberg SE, et al. Rural versus urban pediatric non-accidental trauma: different patients, similar outcomes. BMC Res Notes 2018;11:519.
